# Cost-Effectiveness Threshold for Healthcare and Safety: Justification and Quantification

**DOI:** 10.1101/2021.04.05.21254927

**Authors:** Moshe Yanovskiy, Ori Nissim Levy, Yair Y. Shaki, Avi Zigdon, Yehoshua Socol

## Abstract

Every public expenditure, including saving lives or extending life expectancy of particular persons (target population), has unwanted but unavoidable side effects of statistical shortening of life expectancy of the general public. Therefore, cost-effectiveness analysis in making decisions regarding health and safety is an ethical necessity. We report here cost-effectiveness estimation based on comparison of three independent methods: (1) by analyzing salaries in risky occupations, (2) by assuming that people value their lives twice more than the wealth they earn, and (3) by comparing with the U.S. current legal practice. To the best of our knowledge, nobody applied method (2) to cost-effectiveness analysis. Our result is that the cost-effectiveness threshold for the developed countries is about US$60,000 ± 25,000 per life-year (LY), or about 1.0 ± 0.4 GDP (gross domestic product) per capita per LY. Therefore, a sum of not higher than US$85,000 (1.4 GDP per capita) is statistically sufficient to “purchase” an additional year of life – or, alternatively, to “rob” one year of life if taken away. So, 140% GDP per capita per life-year should be considered as the upper limit of prudent expenditure on healthcare and safety. The result is in excellent correspondence with the existing healthcare policies.

**Highlights:**

- Safety consumes resources; resources are limited
- Therefore, excessive safety expenditure claims more life than it saves
- Prudent safety expenditure is up to 140% GDP per capita per life-year

## 1. Introduction

Investment in health care, technological safety measures, safety and environment-protection regulation decrease risks and extend lives, yet increase the economic burden (Guasoni, & Huang, 2019; Bryndin, 2019; Ma, 2016), which is of paramount importance for low- and middle-income countries. Thus, resource restrictions are expected, and countries will be forced to prioritize medical investments (Stenberg et al., 2017).

Associating human life with monetary value is psychologically difficult. However, a cost-effectiveness analysis in health and safety decision-making is unavoidable (Gold et al. 1996, Viscusi and Aldy 2003, Neumann and Sanders 2017). Cost-effectiveness analysis is routinely performed in health policy, though decisions are rarely, if ever, made based on cost-effectiveness only (Neumann and Sanders 2017). Not only health policymakers but also medical practitioners can no longer consider only benefits and side effects but should also be aware of the associated cost (Bonis and Wong 2019).

Ethical justification of cost-effectiveness analysis for lifesaving and life-extending measures is rarely discussed – it is usually stated that each dollar invested in a potentially life-saving program (for example, cancer prevention) is not invested in another potentially life-saving program (for example, cancer treatment). However, it does not address the frequent claim of ALARA (As Low as Reasonably Achievable) principle proponents: money spent on safety measures (e.g., by private agents due to safety regulation) would not be invested in alternative life-extending projects (HSE, 2001). So, the ethical justification of cost-effectiveness analysis remains an open question.

The rest of this paper is organized as follows. In sect. 2, we address the basic ethical question: why, though the value of human life is the highest value, life extension by means of healthcare and safety should not become a super-goal consuming all the ‘reasonably available’ resources. In sect. 3, by comparing three independent approaches we discuss what amount of money – cost-effectiveness threshold (CET) – is appropriate for life extension. In sect. 4 (Discussion) comparison our results for CET with other estimations is performed, followed by Conclusions.

### 2. Ethical justification of cost-effectiveness threshold for healthcare and safety

The terms ‘cost-effectiveness analysis’ and ‘cost-benefit analysis’ are close. However, cost-benefit analysis places monetary values on health outcomes or life, and therefore raises many ethical objections (Bonis and Wong 2019).

There is an approach that since the value of human life is the highest possible from a social perspective, any risk should be kept ‘as low as reasonably achievable’ – the ALARA principle (HSE 2001), no matter the price, if it is bearable. However, the ALARA approach has an inherent ethical problem: by statistically extending life expectancy of some target population, we statistically reduce life expectancy of the others.

The reason is that well-being and the life expectancy of the modern industrialized society is based on its material basis. Considering an unreasonable extreme, with all resources invested in health care people would just die of hunger. Another extreme – nobody suggests giving up motorized transport though it claims yearly about 35,000 lives in the U.S. alone. In general, each dollar collected as tax (even to be invested in a potentially life-saving program) makes the taxpayer poorer by one dollar. And each dollar spent by a company due to safety regulation decreases by one dollar its profit, with direct impact on the workers’ salaries and shareholders’ income – also making them poorer. And statistically, poorer people have shorter life expectancies.

It has been stated that the impact of socio-economic factors on health is enormous compared to the power of healthcare to counteract these factors. A metaphor for the connection of socio-economic and health parameters is the “New York subway map” (Marmot 2015): From Manhattan to the South Bronx, life expectancy declines by ten years, half a year for every minute on the subway. No medical intervention, either existing or even conceivable, has the same order of magnitude of effect on health.

There are many mechanisms that contribute to the correlation between socio-economic status and life expectancy. Poorer people take higher-risk jobs, they work extra hours stressing their health, they use cheaper, and consequently less safe, products, their lifestyle is less healthy, their diet is less healthy, their health insurance is less comprehensive etc. For example, it has been shown that demand for health insurance is highly elastic (Abraham et al. 2017) – that is, people purchase health coverage plans proportional to their income. Personal experience of one of the authors () illustrates just one of multiple mechanisms leading to lower life expectancy of poorer people: Two of his neighbors lost their lives in two unrelated traffic accidents. Both drove very cheap cars because of their economic status, and both would most probably survive with minor injuries – based on the post factum analysis of the accidents – would each of their cars be just about US$8,000 more expensive. The corresponding loss of life in the two cases was 30 years (50-year-old woman) and 50 years (30-year-old man) – about US$200 per life-year on average.

Not every drop in household income significantly affects health. If the drop is slight, the expenditures on food, medicine and health insurance are usually not affected. The family also tries to preserve the usual vacation patterns, cutting off other outcomes. However, unemployment or a significant reduction in business income will undoubtedly force most affected households to reduce their costs, affecting their health.

Such a stepwise (rather than linear) health effect of income reduction at the microlevel (households) does not mean that a small additional burden placed on the society (for example, a tax) does not pose risks to health and life. Small changes in national income mean significant changes in the expenses of particular households (Blundell, Stoker, 2005). For example, even a small increase in company tax can bankrupt a company balancing on the brink of survival.

It should be emphasized that healthcare costs in most developed countries are already a heavy burden on national economy (Chernew 2011; Ehrlich and Yin 2013): 8-12% of GDP in OECD countries, almost 17% in the USA (OECD 2019 table 1.6 p. 33). Not accidentally, shifting a substantial fraction of health expenditures from the medical system to socio-economic targets has been proposed (Berwick 2020). It is very likely that easing safety regulations and reducing healthcare programs of questionable efficiency will save more life years than lost without them.

**Table 1.**
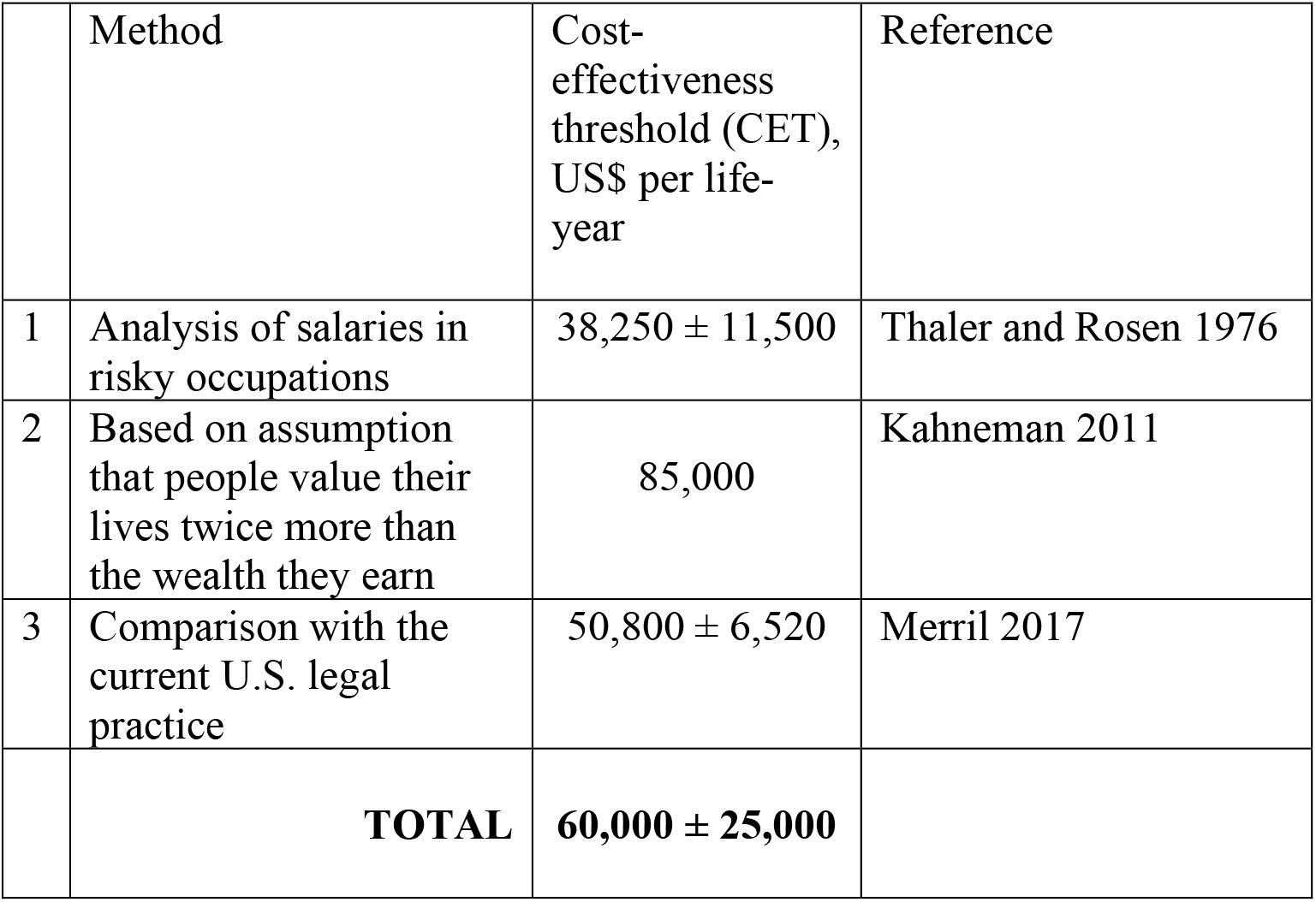
Independent estimations of cost-effectiveness threshold.

| | Method | Cost-effectiveness threshold (CET), US\$ per life-year | Reference |
| --- | --- | --- | --- |
| 1 | Analysis of salaries in risky occupations | $38,250 \pm 11,500$ | Thaler and Rosen 1976 |
| 2 | Based on assumption that people value their lives twice more than the wealth they earn | 85,000 | Kahneman 2011 |
| 3 | Comparison with the current U.S. legal practice | $50,800 \pm 6,520$ | Merril 2017 |
|  | **TOTAL** | **$60,000 \pm 25,000$** |  |

Therefore, every public expenditure, including saving lives or extending life expectancy of particular persons, has unwanted but unavoidable side effect of statistical shortening of life expectancy of the general public. The term *cost-effectiveness threshold* (CET) is widely used among healthcare policymakers; it is assumed that a policy with cost-effectiveness below CET claims more life than it saves.

### 3. Cost-effectiveness threshold

#### 3.1. Quantitative characteristics

The question of decision-making is whether the direct effect, or policy goal (life expectancy extension for the target population), is stronger than the side effect (life expectancy shortening for the general public). If the side effect is stronger than the direct (policy goal), the net result of such policy is the statistical shortening of life (Graham 1995). Such situation has been even characterized by the term “statistical murder” (Graham 1995).

So, quantitative value of public expenditure that statistically leads to loss of one human life (or one life-year) is crucial for decision on efficacy of any potentially-life-saving program.

The term *value of statistical life* (VSL) (Viscusi and Aldy 2003) is in official use by the U.S. government (EPA 2010, Bosworth et al. 2017). While the term VSL probably raises negative connotation of equaling human life to some monetary value, its meaning is just that there is human-life cost for any spending. Essentially, the concept of CET is similar to the concept of VSL in the meaning that both concepts quantitatively assess public expenditure that is worth being invested in safety and other potentially life-expectancy-extending policies and actions.

Another term is *willingness-to-pay* (WTP) generally parallel to CET and VSL, which is often used regarding national health policies. In our opinion, the term WTP, while probably least frequently used, describes most exactly the mechanism behind the assumption that the state should not provide citizens with services (including life-extension services) that are less cost-effective than the citizens themselves are willing to pay for such services (Ngiem et al. 2017). We are going to discuss this mechanism now.

In order to estimate CET, VSL or WTP, one would ideally consider all the reasonable factors – taking risk, safety of products, diet etc. However, this task is formidable due to multiple uncertainties. The most straightforward practical approach to determine CET is probably to estimate how people themselves value their lives in monetary terms. While accepting certain (or even very probable) death for money is unacceptable, taking small risks for money is routine: every profession is associated with some risk and there are professions that are riskier than the others (firefighters, construction workers etc.). If on average people are ready to take risk of death with, for example, probability 1/1000 (one of a thousand) for $1500, then VSL should be estimated as $1.5 million. Namely, to get $1.5 million of earnings, 1000 people on average will take the risk 1/1000 to die, and one on average will die. Though people take their risks voluntary, the net effect is that public expenditure of one VSL statistically claims one human life. The above method of valuing life was proposed by Adam Smith more than two centuries ago and has been used since then in economic analysis as well as in legal practice (Viscusi 2000). Therefore, reasonable value of CET per statistical life saved should be set about the value of VSL. It is not about monetary value of life. It is about extending life population-wide.

#### 3.2. Estimating CET value

Our further analysis is based on three independent estimations: (1) analysis of salaries in risky occupations, (2) analysis based on assumption that people value their lives twice more than the wealth they earn, and (3) comparison with the current U.S. legal practice.

1. In 1976, Thaler (later awarded the Nobel Prize in Economics for 2017) and Rosen analyzed salaries in different occupations and compared the salaries with risk (mortality). They estimated VSL to be $200,000 ± 60,000 in 1967 dollars (Thaler and Rosen 1976). In 2019 dollars, the above estimation corresponds to $1.53M±0.46M based on Consumer Prices Index (https://www.measuringworth.com/datasets/uscpi). In order to perform cost-effectiveness analysis of evacuation we need not only VSL, but also value of spending equivalent to statistical loss of *one life-year*. The latter can be obtained by dividing VSL by half of life expectancy – since statistical (accidental) death can occur randomly at any time during the lifespan. For life expectancy of about 80 years, typical for the developed countries, VSL should be divided by 40 years. The above estimation yields therefore CET = $38,250 ± 11,500 per life-year (the accuracy of the numbers, here and below, is certainly spurious; we keep this spurious accuracy till the final averaging).
2. Thaler’s estimation of VSL fits the rule formulated by Daniel Kahneman (Nobel Prize in Economics for 2002) that people quantify potential loss approximately twice as much as potential earning (Kahneman 2011). According to Kahneman, this rule applies not only to money but also to other goods (vacation days, e.g.). In our case, we assume that people value their lives (that they have and can lose) in monetary terms approximately twice as much as the wealth they anticipate to earn. To the best of our knowledge, such estimation of CET has not been performed before. Let us perform the corresponding calculation. Net average life-time income in U.S. can be estimated as GDP per capita minus taxes. The US GDP per capita was about $65,000 in 2003– 2019 (World Bank 2020). The average US taxes are 35%, leading to an average yearly net income of about $42,500. Doubling this sum yields CET = $85,000 per life-year.
3. In the U.S. legal practice, the median settlement compensation for the victims of the 9-11 attack was $1.7M (in 2017 dollars), and median death compensation awarded by jury in 2009-2013 was $2.2M (also in 2017 dollars) (Merril 2017). We can therefore take $1.95M ± 0.25M for the legal practice with corresponding CET estimation of $48,750± 6,250. In 2019 US$, CET = $50,800± 6,520.

It is worth explaining why legal practice is relevant to the determining of CET. Judicial practices in a country with a respected court accumulate a large volume of practical decisions in various situations. The existing judicial practice, which does not cause condemnation or at least wide discussion, gives therefore a good assessment of the public acceptability of the decision from the point of view of society. Therefore, the assessment of the “value of human life” in court to determine compensation (Viscusi, 2000) provides important guidance for CET/VLS/WTP values acceptable by the society.

The CET values are summarized in Table 1. The difference between the three values is about twofold, which seems to us rather modest taking into account the very different estimation methods.

Rigorous statistical averaging the results of the three methods cannot be done due to many unknowns. Instead of inventing mathematical method of questionable applicability, we just take the CET value as the mean value of the three estimations, and the CET range as the range of the values from US$38,250 to US$85,000. Rounding the numbers to a reasonable accuracy yields:

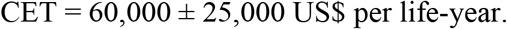

Our estimation was performed on the U.S. data. We can generalize by expressing the result via gross domestic product (GDP) per capita – US$ 65,000 in 2019. Rounding to reasonable accuracy yields:

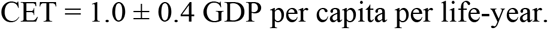

The last number is the single most important result of this study.

## 4. Discussion

Though the value of human life is the highest value for the society, life extension by means of healthcare and safety should not become a super-goal consuming all the reasonably available resources. The reason for this is that every public expenditure, including saving lives or extending life expectancy of particular persons (target population), has unwanted but unavoidable side effect of statistical shortening of life expectancy of the general public.

We find that in the U.S., both people as individuals and the society in general value their lives at about US$60,000 ± 25,000 (1.0 ± 0.4 GDP per capita) per life-year (LY), and therefore a sum of not higher than US$85,000 is statistically sufficient to “purchase” an additional year of life – or, alternatively, to “rob” one year of life if taken away. So, 85,000 US$/LY should be considered as the upper limit of prudent expenditure on safety.

Our value is in excellent correspondence with the values accepted in the field of public healthcare. It has been estimated that the U.S. national health insurance is associated with an average cost-effectiveness of about US$ 50,000 per QALY (quality-adjusted life year) gained (Nghiem et al. 2017). In the United Kingdom, National Institute for Health and Clinical Excellence (NICE) adopted a cost effectiveness threshold range of $40,000 to $60,000 per QALY (Appleby et al. 2007). The review of Bonis and Wong (2017) estimates healthcare CET as US$50,000 to $100,000 per QALY. All these numbers are in excellent correspondence with our estimation.

Our value for CET was derived from three independent evaluations: by analyzing salaries in risky occupations (Thaler and Rosen 1976), by assuming that people value their lives twice more than the wealth they earn (Kahneman 2011), by comparing with the U.S. current legal practice (Merril 2017). Very different numbers can be found in literature; different government agencies estimate VSL (value of statistical life) to justify their policies. Their estimates should be viewed with extreme caution. First, methodologically there are multiple sources of bias, usually towards overestimating VSL (Bosworth et al. 2017).

Second and more important: Any government agency estimating VSL faces explicit conflicts of interest: the higher the VSL, the better the outcome of cost-effectiveness analysis for any proposed policy.

Consider, for example, a policy with a price tag $1000M (one billion dollars). If VSL is $1.5M, such a policy is justified if it saves about 700 people; however, if VSL is $10M – saving 100 people (that is, being 1/7 as effective) is enough for a positive cost-effectiveness judgement. To illustrate the problems with government-estimated VSL we mention that from about 1995 to about 2015, VSL estimates by three U.S. agencies – Department of Agriculture, Food and Drug Administration and Environmental Protection Agency – went up from $2-4M to $9-10M (Merril 2017); VSL estimations of up to US$70 million have been cited in the literature (Viscusi 1994).

Because of high volatility of VSL values reported by different government agencies, and because of the explicit conflict of interest in their VSL estimation, it seems to us prudent to use our value for the cost-effectiveness threshold (CET).

As an example, let us apply CET analysis to the COVID-19 crisis management in Israel. It can be estimated that the direct economic cost of the lockdowns in 2020-21 was about US$ 30 billion, while the Israeli population is about 9.3 million and GDP per capita is about US$ 45,000 (see Appendix). Dividing 30 billion by (1.4×45,000) yields about 500,000 life-years lost; the last number is equivalent, e.g., to life loss due to cancer during 5-year period in Israel. The discussion of whether the above human cost was justified is beyond the scope of this paper.

## Conclusions

Every public expenditure, including saving lives or extending life expectancy of particular persons (target population), has unwanted but unavoidable side effect of statistical shortening of life expectancy of the general public. Therefore, cost-effectiveness considerations in healthcare and safety are not technical, but ethical necessity. Population-wide, a life-extending policy with cost-effectiveness below cost-effectiveness threshold (CET) claims more life than it saves.

Because of high volatility of CET values reported by different government agencies, and because of the explicit conflict of interest in their estimations, it seems to us prudent to use our value for CET, derived by three independent methods and consistent with the current health policies:

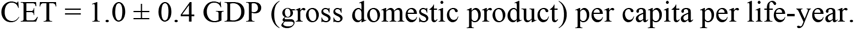

Therefore, 140% GDP per capita per life-year should be considered as the upper limit of prudent expenditure on safety.

## Data Availability

N/A

## Appendix Direct economic effect of lockdowns in Israel

Bank of Israel [1] assessed direct economic losses for four selected periods of restrictions: lockdown I (5.4 billion NIS per week), lockdown II (3.2 billion NIS per week), lockdown III (2.5 billion NIS per week) and “corona-routine” (1.3 billion NIS per week). We applied the above numbers to the following time periods according to the Oxford COVID-19 Government Response Tracker (OxCGRT) prepared by the Blavatnik School of Government [2].

1. Severe restrictions Lockdown I April 1–25: 25 days, 3.5 weeks, 5.4 BNIS/week, 18.9 billion NIS.
2. Significant restrictions
  a. Lockdown II September 25 – October 27: 32 days, 4.5 weeks, 3.2 BNIS/week, 14.4 billion NIS
  b. Lockdown III December 27, 2020 – February 6, 2021 and February 25 – March 1, 2021: 41+5 days, 6.5 weeks, 2.5 BNIS/week, 16.3 billion NIS.
  c. Additional periods of significant restrictions: March 17 – March 30, 2020 (14 days); April 26 – July 12, 2020 (78 days). 92 days, 13 weeks, 2.5 BNIS/week, 32.5 billion NIS.
3. Considerable restrictions “Corona-routine” July 13 – September 24, 2020 (74 days); February 7–24, 2021 (18 days); March 2-31, 2021 (30 days): 122 days, 17.4 weeks, 1.3 BNIS/week, 22.6 billion NIS.

The total cost of lockdowns till March 31, 2021 is therefore 104.3 billion NIS. According to the same document (BOI 2020) the Israeli GDP is about 1400 BNIS. With population of about 9.3 million [3], GDP per capita is about 150,000 NIS. Dividing 104.3 billion by 1.4×150,000 yields about 500,000 life-years lost.

Taking the exchange rate as approximately 3.4 NIS/US$ yields the lockdowns’ cost of $30 billion and GDP per capita of $45,000.

We verified this assessment using data for travel and tourism (T&T) – industry that suffered the most severe blow. During 2019, T&T contributed US$ 22 billion (5.6%) to the Israel’s GDP [4]. The impact of year of missed T&T opportunities is compatible with the above assessment of the overall economic loss.

### Data sources

1. BOI – Bank of Israel. 2020. Press-release Dec 25,2020: Weekly cost of the third lockdown is 2.5 billion NIS per week (in Hebrew). https://www.boi.org.il/he/NewsAndPublications/PressReleases/Pages/25-12-20.aspx
2. Hale, T., Angrist, N., Goldszmidt, R. et al. 2021. A global panel database of pandemic policies (Oxford COVID-19 Government Response Tracker). *Nature Human Behaviour* (2021). https://doi.org/10.1038/s41562-021-01079-8, Tables: https://www.nature.com/articles/s41562-021-01079-8/tables/1
3. Central Bureau of Statistics (Israel). 2020. Media Release: Population of Israel on the Eve of 2021 https://www.cbs.gov.il/en/mediarelease/Pages/2020/Population-of-Israel-on-the-Eve-of-2021.aspx
4. WTTC – World Travel and Tourism Council. 2020. Economic Impact Reports. https://wttc.org/Research/Economic-Impact

## Acknowledgements

The authors wish to thank Prof. Avi Caspi (Jerusalem College of Technology – JCT) for his encouragement of this work. We would like to thank Prof. Shlomo Engelberg (JCT) and Prof. Eli Sloutskin (Bar Ilan University) for thorough reading the manuscript and suggesting many important improvements. We also wish to thank Dr. Moti Brill (Nuclear Research Center Negev, ret.), Prof. Noah Dana-Picard (JCT), Prof. Ludwik Dobrzyński (National Centre for Nuclear Research, Poland), Prof. Marek Janiak (Military Institute of Hygiene and Epidemiology, Poland), Dr. Efraim Laor (Holon Institute of Technology), Prof. Michael Shapiro (Technion – Israel Institute of Technology), Dr. Barak Tavron (Israel Electric Corporation), and Prof. Alexander Vaiserman (Institute of Gerontology, Kiev, Ukraine) for fruitful discussions and constructive criticism.

## References

Abraham J, Drake C, Sacks DW, Simon K. 2017. Demand for health insurance marketplace plans was highly elastic in 2014–2015. Economics Letters 159 (2017) 69–73

Appleby J, Devlin N, Parkin D. 2007. NICE’s cost effectiveness threshold. How high should it be? BMJ 2007; 335

Berwick DM. 2020. The Moral Determinants of Health. JAMA 324(3):225–226. DOI: 10.1001/jama.2020.11129

Blundell R., Stoker T.M. 2005. Heterogeneity and Aggregation. Journal of Economic Literature, 43 (2): 347–391.

Bonis P.A.L and Wong J.B. 2019. A short primer on cost-effectiveness analysis, UpToDate®. Alphen aan den Rijn: Wolters Kluwer. https://www.uptodate.com/contents/a-short-primer-on-cost-effectiveness-analysis Updated: Nov 13, 2019. Accessed July. 1, 2020.

Bosworth R.C, Hunter A, Kibria A, The Value of a Statistical Life: Economics and Politics. Logan, UT: Strata, 2017. https://strata.org/pdf/2017/vsl-full-report.pdf Accessed Jan. 1, 2020.

Chernew M. 2010/2011. Health Care Spending Growth: Can We Avoid Fiscal Armageddon? Inquiry 47(4):285–295.

Domb C. 1995. Common Sense, Scientific Method and Accident Prevention. Jerusalem, Nebenzahl Institute for Human Safety and Accident Prevention.

Ehrlich I., Yin Y. 2013. Equilibrium Health Spending and Population Aging in a Model of Endogenous Growth: Will the GDP Share of Health Spending Keep Rising? Journal of Human Capital, 7(4): 411–447.

EPA (U.S. Environmental Protection Agency) 2010. Valuing Mortality Risk Reductions for Environmental Policy: A White Paper. December 10, 2010 https://www.epa.gov/sites/production/files/2017-08/documents/ee-0563-1.pdf Accessed Jan. 1, 2020.

Gold M.R., J.E Siegel, L.B. Russell, and M.C. Weinstein (eds). Cost-effectiveness in Health and Medicine. New York: Oxford University Press, 1996

Graham JD. 1995. Comparing Opportunities to Reduce Health Risks: Toxin Control, Medicine and Injury Prevention. NCPA Policy Report No. 192, National Center for Policy Analysis, Dallas TX.

Guasoni, P., & Huang, Y. J. (2019). Consumption, investment and healthcare with aging. Finance and Stochastics, 23(2), 313–358. https://doi.org/10.1007/s00780-019-00383-6

HSE (Health and Safety Executive), 2001. Reducing Risk, Protecting People: HSE’s Decision Making Process, Her Majesty’s Stationery Office, Norwich NR3 1BQ.

Kahneman D. 2011. Thinking, Fast and Slow, NY, Farrar, Straus and Giroux

Korn G.A. and Korn T.M. 1968. Mathematical Handbook for Scientists and Engineers. NY, McGraw Hill, §19.6-6

Ma, Y., Zhao, Q., & Xi, M. (2016). Decision-makings in safety investment: An opportunity cost perspective. Safety science, 83, 31–39. https://doi.org/10.1016/j.ssci.2015.11.008

Marmot M. 2015. The Health Gap: The Challenge of an Unequal World. London: Bloomsbury, ISBN: 9781408857991.

Merrill D. 2017. No One Values Your Life More Than the Federal Government. October 19, 2017. https://www.bloomberg.com/graphics/2017-value-of-life/ Accessed Jan. 1, 2020.

Neumann P.J and Sanders G.D. Cost-Effectiveness Analysis 2.0. N Engl J Med 2017; 376:203–205

Nghiem S, Graves N, Barnett A, Haden C (2017) Cost-effectiveness of national health insurance programs in high-income countries: A systematic review. PLoS ONE 12(12):e0189173. https://doi.org/10.1371/journal.pone.0189173 Accessed Jan. 1, 2020.

OECD 2019. Health at a Glance 2019: OECD Indicators, Paris: OECD Publishing, https://doi.org/10.1787/4dd50c09-en.

Stenberg, K., Hanssen, O., Edejer, T. T. T., Bertram, M., Brindley, C., Meshreky, A., … & Soucat, A. (2017). Financing transformative health systems towards achievement of the health Sustainable Development Goals: a model for projected resource needs in 67 low-income and middle-income countries. The Lancet Global Health, 5(9), e875–e887. https://doi.org/10.1016/S2214-109X(17)30263-2

Thaler R and Rosen S. 1976. The Value of Saving a Life: Evidence from the Labor Market. In: Nestor E. Terleckyj (Ed.). Household Production and Consumption, pp. 265-302. NBER 1976

Viscusi W.K. 1994. Mortality Effects of Regulatory Costs and Policy Evaluation Criteria. The RAND Journal of Economics, 25(1): 94-109. Available at: http://www.jstor.org/stable/2555855 Accessed Jan. 1, 2020.

Viscusi W.K. 2000. The Value of Life in Legal Contexts: Survey and Critique. American Law and Economics Review, 2(1):195–222.

Viscusi W.K. and Aldy J.E. 2003. The Value of a Statistical Life: A Critical Review of Market Estimates throughout the World, Journal of Risk and Uncertainty, Springer 27(1): 5–76.

World Bank 2020. GDP per capita – United States. https://data.worldbank.org/indicator/NY.GDP.PCAP.CD?locations=US Accessed July 1, 2020.

